# Exploring perceptions and attitudes toward pain in surgical trauma patients in a public hospital in Guatemala: a qualitative study

**DOI:** 10.64898/2026.11.11.26367874

**Authors:** Vikas Mahalingam, Ryan Turner, Karin Zacarias, Sindy Ramirez, Colby G. Simmons, Jakob E. Gamboa

## Abstract

Pain is nearly universal following traumatic injuries. However, pain perception, expression, and coping may be shaped by biological, psychological, social, and cultural factors that remain poorly characterized among underrepresented populations. This study explored experiences, attitudes, and expectations toward pain among hospitalized trauma patients at a public hospital in Guatemala.

We conducted a qualitative study using semi-structured interviews with consenting adults admitted to the orthopedic trauma service at Hospital Nacional de Coatepeque in southwestern Guatemala from May to June 2025. Interviews explored pain experiences, coping strategies, interactions with healthcare providers, and expectations for pain management. Interviews were conducted in Spanish, audio-recorded, transcribed verbatim, translated into English by bilingual researchers, and analyzed using inductive thematic analysis.

Five interrelated themes were identified: 1) lived experience and trajectory of pain; 2) coping, adaptation, and psychological endurance; 3) spirituality, meaning-making, and moral framing of suffering; 4) pain management and interactions with the healthcare system; and 5) social context, responsibilities, and life disruptions. Participants described severe early pain accompanied by physical, emotional, and functional distress that generally improved over time. Psychological coping, faith, and family responsibilities were central to resilience and recovery. Notably, patients often reported acceptable or improving numerical pain scores despite narratives describing substantial physical, emotional, and functional suffering, suggesting that standardized pain scales may not fully capture lived experiences.

Trauma patients in Guatemala described pain as a multidimensional experience shaped by physical, psychological, spiritual, and sociocultural factors. Despite severe pain, patients demonstrated resilience, often accepting pain as a temporary part of healing while prioritizing recovery and return to family responsibilities. The discordance between numerical pain scores and narratives of suffering underscores limitations of pain scores as standalone measures. These findings support culturally informed, patient-centered strategies that integrate pharmacologic treatment with attention to psychological, spiritual, and social dimensions of recovery.

## Introduction

Pain after trauma is a complex experience that is shaped not only by injury severity but also by communication, access, and sociocultural context [1–3]. Variability in clinical practice and differences in patient-centered approaches can influence how pain is assessed and managed, particularly in high-acuity environments such as trauma and critical care [4,5]. Trauma presents unique challenges, as time-sensitive decision-making and high patient acuity may limit opportunities for comprehensive pain evaluation and patient input for comprehensive assessment [6]. Therefore, the ability to understand how patients perceive, communicate, and prioritize pain within these unique settings is essential to improving clinical outcomes and the quality of patient-centered care and communication.

The perception of pain can be greatly influenced by cultural and contextual factors, which shape expectations, experiences, and coping strategies. Available literature highlights that individual differences in neurological processing, prior experiences, and emotional context contribute to variability in pain perception and response to treatment [7,8]. Cultural beliefs about pain expression, stigma, and acceptable coping mechanisms may further influence whether and how patients report pain, while health literacy can affect their ability to communicate symptoms effectively [7,9]. Facilitating patient agency and empowerment may be a crucial factor for improved perceptions and outcomes [10]. These considerations underscore the importance of culturally responsive and contextually aware approaches to addressing pain.

The challenges of understanding and addressing may be magnified in rural and underserved populations, where access to healthcare resources is often more limited. Factors such as fewer healthcare providers, long travel distances, and limited access to specialized care can all affect how pain is managed, while local culture, norms and community contexts may influence expectations around perceived pain level, suffering, resilience, and behaviors around seeking [11–13]. Examining perceptions of pain among trauma patients in underserved populations is critical to identifying gaps in care and informing more equitable, patient-centered approaches to pain assessment and treatment. The aim of this study is to examine patient-reported experiences with acute pain among underserved populations admitted with traumatic injuries in a resource-limited, public hospital in Guatemala.

## Methods

### Study Design

This qualitative descriptive study explored patients’ experiences, perceptions, and attitudes regarding pain and pain management during hospitalization at a public hospital in Guatemala. Semi-structured interviews were conducted with patients hospitalized for traumatic injuries at the Hospital Nacional de Coatepeque (HNC) from May 19, 2025 to June 6, 2025. We used an inductive qualitative approach, which enables an in-depth understanding of patients’ experiences and perspectives from underrepresented populations which have not been characterized previously [14]. The COREQ (consolidated criteria for reporting qualitative research) was used as a guideline for reporting the study [15]. The complete COREQ checklist is provided in S1 checklist.

### Study Setting and Participants

The Hospital Nacional de Coatepeque is a public, first-referral hospital in the town of Coatepeque in southwestern Guatemala. The hospital serves a patient population of an estimated 150,000, which includes many rural, agricultural communities in the southwest region who experience among the highest rates of poverty and worst health outcomes in the country [16,17]. The region experiences a high burden of trauma, primarily from motor vehicle accidents or work-related injuries. HNC contains two orthopedic trauma wards separated by gender each with approximately 20 beds.

Adult individuals, age ≥18, who were admitted to the orthopedic trauma service during the study period and consented to participate were eligible. Exclusion criteria consisted of refusal to participate, altered mental status, or inability to consent. Purposive sampling was performed through nursing staff screening and recommendations of all eligible admitted patients within their respective services. The researchers intentionally sought individuals with varying ages, genders, and injuries to capture broad representations of experiences [18]. In this study, theoretical saturation was achieved with a final sample of 20 patients. This refers to the point when interviews yielded no substantially new information or themes [19]. No patients refused to participate.

### Data Collection

Semi-structured interviews were conducted in person in the preferred Spanish language of the region with a researcher (JG) who is a male, American Board-Certified Anesthesiologist, certified medical interpreter, and qualified professional bilingual speaker with experience in qualitative and survey research. The interviewer had no prior relationship with the participants and was not a provider of direct clinical care within the study setting, reducing the likelihood of power dynamics or response bias. An interview guide was used (S1 Text) and was developed based on literature and expert review and team consensus. Two native Spanish speakers reviewed the interview guide, and revisions were made to ensure local linguistic clarity and comprehension. Ad-hoc questions were asked during the interview to explore further meaning of responses. Additionally, basic demographic information and pain and injury characteristics were queried using a Spanish Wong-Baker FACES® Pain Rating Scale. The Wong-Baker FACES® Pain Rating Scale is a self-reported pain intensity measure in which patients select the face and corresponding score from 0 to 10 that best represents how much pain they are experiencing [20]. This validated instrument was used due to its visual graphics to reduce barriers related to literacy.

Eligible participants were approached in person during their hospitalization by the interviewer who introduced himself and his role, described the study purpose, and obtained verbal informed consent prior to enrollment. Verbal informed consent was approved by both ethical review boards in lieu of written consent given the minimal-risk nature of the study and the absence of identifiable information collected. Consent was obtained prior to each interview and documented within the audio recording. Due to the inability to transport or remove injured patients from the trauma service, interviews were performed discreetly at bedside with the patients’ permission, in a manner to promote privacy and ensure that nearby patients could not overhear the conversation. Other parties that may have been inadvertently present (within earshot) include hospital staff and neighboring patients.

Three pilot interviews were performed to test the interview guide and ensure comprehension of questions, and minor modifications were implemented for subsequent interviews. During interviews, repeating information was noted, and saturation was reached, with two additional interviews conducted for confirmation. The interviews ranged from 8 to 46 minutes in duration. Audio recordings were transcribed verbatim in Spanish and subsequently translated into English by language-proficient members of the research team. To preserve meaning and contextual accuracy, translations were reviewed collaboratively by bilingual researchers familiar with regional linguistic nuances and clinical terminology. De-identified transcripts were stored securely and used for analysis. No field notes were made during or after interviews.

### Data Analysis

Data were analyzed using thematic analysis following the six-phase approach described by Braun and Clarke, an iterative method where reviewers familiarize themselves with the text to code data and identify themes [21]. An inductive analytic strategy was employed to allow themes to emerge directly from participant narratives rather than from predetermined theoretical frameworks. Two researchers independently reviewed and coded transcripts. Codes were refined iteratively through team discussion and consensus and then organized into categories for theme abstraction. Codes were allowed to map onto more than one category where overlapping concepts existed (e.g., a code capturing prayer-based coping mapped to both “Coping and acceptance with pain” and “Belief about the nature of pain”), reflecting the interrelated nature of the pain experience, and allowing for the inclusion of minor themes which later consolidated into the paper’s five final themes. Themes were first generated independently and then discussed as a team; multiple rounds of discussions were conducted until consensus of theme labels was reached. In-vivo quotes and excerpts were also highlighted to illustrated concepts and themes from the text and preserve participant voice. Dedoose qualitative software version 10.09 (SocioCultural Research Consultants, LLC) was used to manage and organize codes. The full coding framework is provided in S1 Table.

### Trustworthiness

To enhance trustworthiness and analytic rigor, coding and theme development were conducted collaboratively by multiple investigators. Regular team discussions were used to review coding consistency, resolve discrepancies, and refine thematic interpretations. Triangulation was performed through multiple methods, including collaborative review and interpretation of interview data. Peer debriefing was conducted with local Guatemalan research team members to improve contextual interpretation, cultural validity, and credibility of findings. Due to limitations in patient follow-up and unreliable communication after hospital discharge, researchers were unable to conduct participant member checking with interviewees after data collection or discuss final themes directly with participants. Findings will be shared with local clinicians and hospital leadership at Hospital Nacional de Coatepeque to identify feasible, locally appropriate strategies to improve pain assessment and patient-centered pain management.

### Ethical Considerations

Ethical review exemption was obtained by the Colorado Multiple Institutional Review Board (COMIRB #25-0713) and the Hospital Nacional de Coatepeque Committee for Bioethics and Scientific Investigation (Comité de Bioética e Investigación Científica) due to minimal risk posed to participants. Participants provided verbal informed consent, which is standard practice at the hospital, due to the low literacy rates among the patient populations served at this hospital. All participants were informed of the study’s aims, procedures, and were assured that participation would not affect their care, data would remain confidential, and that they could withdraw or refuse to answer questions; patients expressed their understanding of the study protocol and were prompted to ask any questions prior to opting for participation. All transcript data were stored without patient identifiers.

## Results

Twenty patients participated in semi-structured interviews (40% female; mean age 43 years). Participants were interviewed an average of 9.6 days after injury and reported a range of injury mechanisms, including motorbike and car collisions, falls, and pedestrian injuries predominantly involving the lower and upper extremities (Table 1).

**Table 1.**
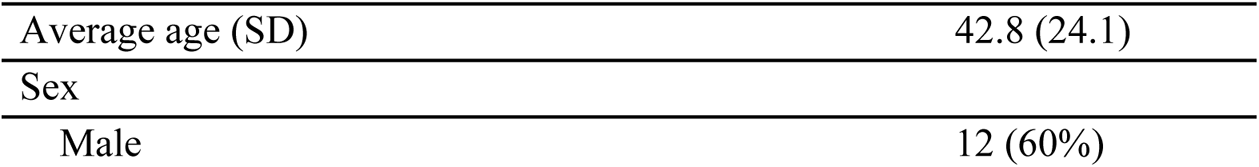
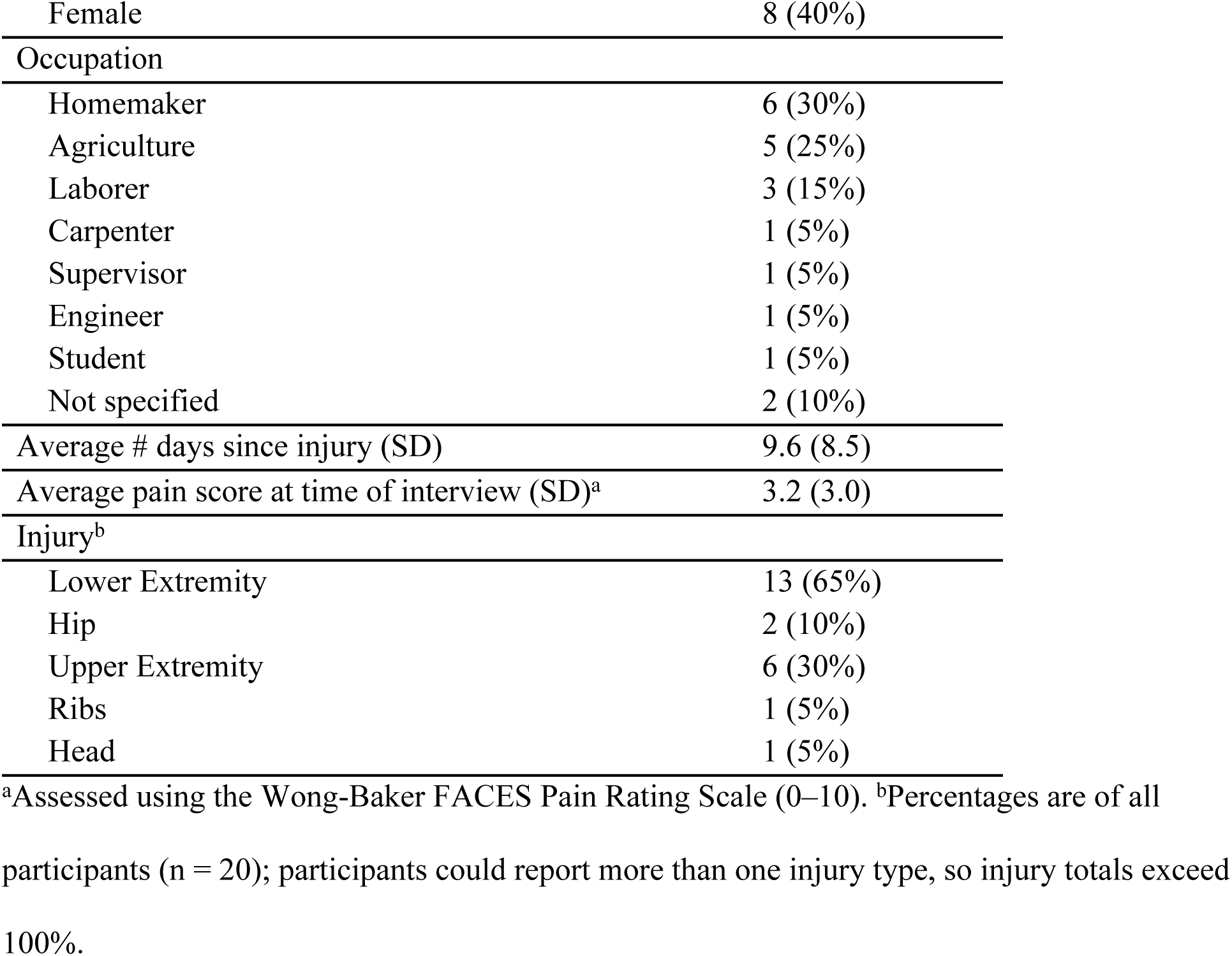
Interview participant characteristics.

Themes were derived following data coding and analysis. Thematic analysis generated five major themes: (1) the lived experience and trajectory of pain, (2) coping, adaptation, and psychological endurance, (3) spirituality, meaning-making, and moral framing of suffering, (4) pain management and interactions with the healthcare system, and (5) social context, responsibilities, and life disruption. Themes were interrelated, reflecting the complexity of the pain experience in this population.

### Theme 1: The Lived Experience and Trajectory of Pain

This theme was the most richly coded in the dataset (n = 94) and encompassed the full arc of patients’ physical and emotional encounters with pain, from initial injury through hospitalization. Participants consistently described severe pain at the onset of their injuries, with many characterizing the experience as overwhelming or unbearable. Pain affected multiple physiologic domains: disrupting sleep, limiting appetite, causing headaches, and restricting mobility. This was frequently accompanied by emotional distress, anxiety, and psychological trauma. Several participants described struggling with helplessness and discouragement in response to their pain, particularly in the context of prolonged immobility or extended hospital stays.

> “Yes, I’m in a lot of pain.. the pain is suffocating. Very intense.” (P5)

> “Sometimes a 10 out of 10. Well, the truth is, I’m only somewhat relaxed today because yesterday I spent about an hour crying from the pain. Because I couldn’t stand it anymore.” (P20)

> “I don’t cry so much because it hurts. I can bear the pain. I take a pill and that’s it. But the helplessness I feel, the uselessness I feel now, of not being able to do this or that.” (P9)

Despite this severity, a notable trajectory of improvement emerged across narratives. Most participants reported meaningful reductions in pain over the course of their admission, attributing relief to medications, surgical intervention, rest, and the passage of time. Participants spoke of viewing surgery as a step toward recovery and described hopefulness for pain reduction following their procedures. Many expressed a practical and at times accepting orientation toward pain thereby acknowledging it as an expected part of the healing process while still clearly identifying it as burdensome. Interesting variation was observed in how patients characterized their pain numerically: some reported discordance between numerical rating scale scores and their verbal descriptions, occasionally endorsing “no pain” while describing ongoing discomfort, suggesting cultural or contextual factors in pain self-reporting.

> “I’m calm. I feel that after the operation, there’s going to be horrible pain after the operation. But I feel that that pain will be part of my recovery.” (P16)

> “And the pain? I think… It’ll be better soon. It’ll be more comfortable, I’m sure” (P14)

> “You just have to be patient with what’s happening… I have to be patient and wait because you can’t… you go somewhere else, and you won’t get better. Yes, this way of improving or you won’t improve” (P12)

### Theme 2: Coping, Adaptation, and Psychological Endurance

Participants drew on a wide and inventive repertoire of coping strategies to manage their pain experience. Non-pharmacologic approaches included repositioning and strategic immobility, breathing exercises, distraction through entertainment, music, reading, and mobile technology, as well as seeking social connection through conversation with family members, other patients, or hospital staff. Several participants described deriving comfort from caring for others in the ward to redirect attention away from their own suffering.

> “I try to close my eyes and fall asleep while the pain passes. And if not, I just breathe. I just have to breathe.” (P8)

> “Well, staying calm. Trying not to relax and stay calm without stressing too much so the pain doesn’t overwhelm me. Yes, not thinking about it too much. Because the more importance you give it, the more it affects the pain.” (P21)

Psychologically, participants employed active mental reframing — recasting pain as temporary, as surmountable, or as a stimulus for strength — as well as more passive strategies such as self-calming and deliberate disengagement from painful sensations. A strong ethic of endurance and stoicism was evident: participants frequently expressed pride in not complaining, described themselves as hardworking and self-sufficient, and spoke of choosing to “get through” their pain rather than expressing it openly. Duty-driven coping was also prominent, with participants citing responsibility to family and a need to return to their roles as motivators for perseverance. Some participants, however, disclosed that the hospital experience altered their mood and emotional state in ways that were difficult to manage, highlighting the limits of adaptive coping under prolonged adversity.

> “Ignoring it, not paying much attention to it because the more you stress, the more pain you feel. It’s better to ignore it, because we’re broken, and that way it’s less painful…As some say, it’s psychological.” (P3)

> “I think that the last thing the pain should take away is the fighting spirit, because the day you lose that, you’re lost.” (P18)

> “As the days go by, if you convince yourself that the pain is mental, its lessening, the pain is decreasing a little” (P14)

### Theme 3: Spirituality, Meaning-Making, and Moral Framing of Suffering

A spiritual dimension of the pain experience was distinctly expressed by many. Participants frequently referenced faith and religion as sources of comfort, resilience, and explanation. Trust in divine help, prayer as a pain-relief strategy, and the belief that God was ultimately in control of their situation were commonly expressed. Many participants described suffering as holding a deeper meaning, whether as a test of character, an occasion for moral reflection, or a catalyst for spiritual growth.

> “I ask God to take away the pain because I cannot handle it, but I can think about Him” (P4)

> “Yes, I pray to God a lot to take away the pain, because He alone is the only one who helps us and He is in the hands of the doctors.” (P7)

Notably, some participants described interpreting their pain through a moral or philosophical lens, reflecting on culpability, the cause of their suffering, and what their experience signified about their lives. This orientation coexisted with a normalization of suffering — a shared understanding that pain was to be expected and even accepted as part of human experience. The concept of mental reframing within a spiritual register was also evident, with participants describing positivity and faith as forces capable of diminishing pain. This theme intersected substantially with Theme 2, as spiritual resources functioned as a primary coping mechanism for many participants.

> “So I believe in Him and I say to Him, ‘Lord, help me to have the strength, to have patience,’ just like you have with all of us, right? Because He has patience with the good and the bad-the temperamental and the non-tempered, right? That’s right. And that gives me strength. That gives me courage” (P15)

> “Trust in God. I know it’s a test. Yes. And I know that I have to get through this, one way or another, because I have to get through this. But more than anything, it’s a test. Sometimes you’re rebellious, and until your ears are pulled… you begin to understand what God wants for your life. Yeah, but I do have faith that I have to get up from this.” (P8)

### Theme 4: Pain Management and Interactions with the Healthcare System

This theme encompassed participants’ experiences seeking, receiving, and evaluating pain treatment within the hospital setting. Overall, participants expressed largely positive views of their medical team and reported satisfaction with the care they received. Trust in providers was a recurrent motif, and many participants described deferring to the medical team’s judgment regarding their treatment. Scheduled pain medications were identified as a primary source of relief, with participants recognizing the role of analgesics in improving comfort and sleep. Injected medications were specifically referenced by some as effective interventions.

> “Well, the nurses gave me an injection and the pain is gone now, so I can sleep better.” (P6)

> “They do a great job and provide excellent care. They provide very good care. They are patient and provide quality care.” (P7)

However, several participants also described gaps and frustrations in their care. Themes of uninformed or insufficient pain management emerged, with some patients reporting incomplete relief from their medications or limited availability of resources. A subset of participants described the need to self-advocate for adequate pain control, requesting medications themselves rather than having pain proactively assessed. Others expressed discomfort discussing their pain openly with providers. Delays in care and prolonged waiting periods, including pre-surgical waiting, were noted as stressors. Participants also described awareness of resource limitations and scarcity in this setting, sometimes adjusting their expectations accordingly. Despite these challenges, the dominant orientation toward the healthcare system was one of gratitude and trust.

> “Because you have to complain to get them to give you medication. Because they know that the medication… they can’t give you one every few minutes, at least.” (P8)

> “Because, as you can see, more patients are coming in, and the ones who are the worst affected are the ones they want to hold on to. Because the space is so small, we have to keep coming in and out.” (P7)

### Theme 5: Social Context, Responsibilities, and Life Disruption

Participants described their pain experience within the broader fabric of their lives and communities. Injury and hospitalization were understood not only as physical events but as disruptions to deeply held social roles and obligations. Many participants were engaged in manual labor occupations, and the inability to work, combined with financial concerns, emerged as a significant source of distress, often compounding the psychological burden of pain itself. The need to return home and resume caregiving responsibilities for family members was frequently cited as a primary motivator for recovery.

> “It’s nice here, but I need to get to my house because I have my children.” (P10)

> “I feel discouraged I’m used to my home, to doing my chores, and I don’t like being in bed… But I get desperate, not doing anything, just lying down, because I’m used to doing everything.” (P15)

Loneliness and the emotional weight of being separated from home and family were prominent, and several participants expressed a strong desire to leave the hospital as soon as possible. Dependency on others, whether for personal care, mobility, or daily tasks, was experienced as difficult by participants who identified strongly with independence and self-sufficiency. Legal concerns related to their injuries were raised by some, adding an additional layer of complexity to their experience. At the same time, social connection functioned as both a source of pain relief and emotional sustenance: family visits, conversation, and a sense of reciprocity with staff and other patients were described as meaningful. Participants also reflected on the relative merits of hospital versus home environments, with some acknowledging that despite the discomfort, hospital care offered access to treatment unavailable at home.

> “I say, it’s true that I’m here in bed, I’m not at home, but I know that by getting better here, that’s the hope. By getting better here, I’ll go home healthy and go back to doing my chores.” (P15)

## Discussion

This study explored the lived experience of pain among adult trauma patients at a public hospital in southwestern Guatemala. Five interrelated themes emerged, reflecting a dynamic pain experience that was simultaneously profoundly physical, emotionally complex, spiritually mediated, and embedded in social roles and obligations. These findings illuminate the biopsychosocial dimensions of pain and importance of culturally responsive, patient-centered pain management in an underrepresented population in resource-limited healthcare settings.

Participants in this study consistently described severe pain at injury onset, with significant emotional distress accompanying physical pain. This finding aligns with existing evidence that trauma-related pain carries psychological as well as nociceptive burden [22]. Despite this initial severity, most participants described a meaningful trajectory of improvement over the course of their hospitalization, which they attributed to outlook, attitude, medications, surgical intervention, rest, and time. The arc from acute overwhelming pain toward cautious optimism mirrors findings from other qualitative studies in surgical populations, in which patients are oriented toward recovery and frame their suffering as temporary and purposeful [10,13].

Notably, participants demonstrated frequent discordance between numerical pain scores and verbal pain descriptions often endorsing “no pain” or minimal scores while simultaneously describing ongoing discomfort, functional limitation, and emotional distress. This finding resonates with literature demonstrating that unidimensional numerical rating scales are affected by a range of social and cultural factors, including concern about being perceived as weak or complaining, trust in providers, and a desire to avoid burdening others [12,23]. The International Association for the Study of Pain has specifically highlighted that numerical rating scale instruments have limited utility in some non-Western and rural populations, where verbal or faces-based rating approaches may better capture the patient experience [24]. Yet, in our study the Wong-Baker faces scale was used with visual and numerical representations of pain but remained inadequate in classifying overall levels of suffering. This may be due to the unique setting in Guatemala, where stoicism and a cultural norm of non-complaint were standard, and where numerical tools likely underestimate the true pain burden. Further investigation is needed to explore these gaps in other similar settings to better understand limitations with unidimensional pain assessments in diverse, underrepresented patient populations.

A central finding was that patients described themselves as active participants in managing their pain rather than passive recipients of treatment. Participants employed numerous cognitive and behavioral coping strategies, including distraction, positive reframing, prayer, emotional regulation, and acceptance to preserve emotional control and progress through recovery. This finding aligns with the broader literature demonstrating that non-pharmacologic coping is a cornerstone of the patient pain experience across setting and cultural environment [10,13]. Of note, the emphasis participants placed on self-directed recovery may partly reflect the rehabilitation context at HNC, where formal physical therapy is not available and nursing staff face significant staffing constraints. In this setting, patients and families often assume greater responsibility for mobility and rehabilitation, potentially reinforcing the sense of ownership and personal agency that characterized participants’ approaches to pain and recovery.

Stoicism and endurance were evident among participants, who frequently expressed pride in not complaining, an explicit reluctance to express pain openly, and a framing of psychological endurance as not merely adaptive but morally virtuous. This orientation has been anthropologically documented with the cultural notion of “simpatía” which beckons values of stoicism and fatalism in the ultimate goal of maintaining relational harmony [25,26]. This prioritization of social agreeability has been shown to shape pain communication and help-seeking behavior [27]. Research consistently demonstrates that in diaspora populations within the United States, diasporic patients are less likely to request pain medication, even when in significant distress [28]. While this stoicism may reflect genuine resilience, it also risks contributing to underreporting and undertreatment, highlighting the need for clinicians to recognize cultural influences on pain expression [29].

Spirituality emerged as another prominent source of resilience and was closely intertwined with psychological coping rather than functioning as a separate construct. The robust role of spirituality in participants’ pain experience is consistent with the religious and cultural landscape of Guatemala, where Catholicism and Evangelical Christianity are deeply embedded in daily life and community identity [30]. Prayer was widely described as an active and practical pain-relief strategy, and trust in divine will provided a framework for accepting and enduring suffering. This finding is strongly supported by evidence from Latin American patient populations more broadly. A multicenter study including Guatemalan patients receiving palliative care for advanced cancer found that 97% identified as spiritual and 89% as religious, and that spirituality functioned as a significant coping resource [31]. Notably, spiritual needs frequently went unmet by medical teams. Similarly, research on pain and spirituality in Brazilian cancer patients demonstrated that spiritual well-being was an adaptive coping strategy and a predictor of improved quality of life [32]. These parallels suggest that the value of spirituality in our trauma population is not incidental but reflects a durable cultural and psychological orientation toward suffering that extends across disease contexts. Acknowledging patients’ spiritual beliefs may therefore represent an important component of culturally responsive perioperative care and improved perceived health outcomes in this patient population.

Participants expressed largely positive views of their providers and high levels of trust in medical care. This is a finding consistent with evidence from other qualitative studies in surgical populations [10]. Pain medications were viewed as effective sources of relief, though analgesia was often described as reactive, with the need to self-advocate when pain was severe. This observation likely reflects both resource limitations and cultural expectations surrounding pain expression and acceptance. Structural gaps in systematic and proactive pain assessment have been described in other resource-limited surgical settings [12,13]. Additional factors, such as the tendency of patients to defer to medical authority in many global healthcare contexts and the adjustment of expectations in response to perceived resource constraints may compound these challenges and result in underreporting and undertreatment. Rogger et al. [7] propose that a holistic and culturally sensitive approach that includes awareness, communication, and training is likely to improve pain management outcomes in diverse and underserved patient populations. The current findings suggest that systematic pain assessment protocols and patient education regarding the need and appropriateness of advocating for pain relief could improve the quality of care in this setting.

Pain was also experienced within a broader social context that frequently outweighed pain itself. Many participants expressed greater concern about prolonged hospitalization, inability to work, financial hardship, dependence on others, loneliness, or returning home to care for family than about pain intensity alone. Manual labor occupations, which were predominant in this population, created a particular vulnerability, as injury directly threatens economic livelihood and caregiver responsibilities. These findings reinforce previous literature demonstrating that recovery after traumatic injury is embedded within patients’ social roles and economic responsibilities, particularly among rural and underserved populations globally [11–13]. Thus, recovery was often framed not merely as a personal goal but as a social obligation. Understanding this orientation may help clinicians better communicate treatment timelines and recovery expectations in ways that are meaningful and motivating for select patient groups. Loneliness and separation from family during hospitalization were identified as significant sources of distress with occasional recognition as an opportunity for social connection. Family visits, conversation with staff and fellow patients also functioned as a meaningful source of pain relief and emotional sustenance. This reflects the broader literature on social modulation of pain perception and suggests that simple interventions supporting social connection during hospitalization such as facilitated family visits, communal ward interaction, and provider engagement may have genuine analgesic as well as psychological value [33,34].

### Implications for Clinical Practice and Policy

Multiple practice-relevant implications arise from these findings. First, standard numerical pain rating tools may underestimate pain burden in this setting due to stoicism, cultural norms of acceptance and communication of pain, and limitations of scale-based assessments. Supplementary qualitative assessment incorporating questions regarding function, sleep, mood, and daily activities may yield a more complete assessment. Second, proactive and systematic pain assessment protocols are needed to ensure adequate pain treatment in a setting where patients are unlikely to volunteer reports of uncontrolled pain. Educational interventions as well as provider-based tools that normalize advocacy for pain relief may help close this gap. Third, the salience of spirituality, family, and social role as coping resources suggests that pain management in this setting should be broadly conceived. Brief acknowledgment of spiritual orientation, facilitating family involvement where feasible, and contextualizing treatment within patients’ recovery goals, including the goal of returning to caregiving and occupational roles, may enhance both engagement and outcomes. Finally, the resource constraints described by participants such as limited analgesic availability, scarcity of providers, and delays in care may represent systemic barriers that qualitative research alone cannot resolve. Advocacy for adequate analgesic formulary and staffing at facilities like HNC is a necessary step to enhancing patient-centered analgesia.

### Limitations

This study has several limitations. The sample was drawn from a single hospital during a single data collection period, which limits generalizability to other regions or settings within Guatemala. Cultural nuances of Guatemalan pain perceptions limit generalizability across Latin America. Although theoretical saturation was reached, the limited sample size of participants reflects the exploratory nature of the study, and larger or longitudinal studies would be needed to fully characterize the trajectory of pain experiences over time and following discharge. Due to logistical constraints, participant validation through member checking after data collection was not feasible due to the unique constraints with communication and patient follow up, which limits the confirmatory depth of the findings. Interviews were conducted at the bedside in a multi-patient ward, which, despite precautions to ensure privacy, may have introduced social desirability bias in participants’ responses. This is a concern consistent with the observed cultural norms around faith, stoicism, and acceptance. Finally, the study captured a Spanish-speaking population; the experiences of indigenous-language speakers in this country, who face additional barriers related to language and healthcare access, are not represented and warrant dedicated investigation.

## Conclusion

Trauma patients in Guatemala described pain as a multidimensional experience influenced by physical recovery, psychological adaptation, spirituality, healthcare interactions, and social responsibilities. Although severe pain was common, patients demonstrated remarkable resilience and frequently accepted pain as a temporary component of healing while prioritizing restoration of function and return to family and work. Patients often minimized pain numerically despite describing substantial suffering, suggesting that pain scores alone may inadequately capture the lived experience of pain in this setting. These findings highlight opportunities to strengthen patient-centered pain management by integrating pharmacologic treatment with culturally informed approaches that acknowledge the psychological, spiritual, and social dimensions of recovery.

## Data Availability

The data underlying this study consist of audio recordings and interview transcripts from 20 hospitalized trauma patients at a single public hospital. Participants provided verbal consent to participate in the research but did not consent to public release of their interview data. Given the small sample size and the specific clinical and demographic details contained in the transcripts, de-identified data could still permit indirect identification of participants. For these reasons, the Colorado Multiple Institutional Review Board and the Comité de Bioética e Investigación Científica of Hospital Nacional de Coatepeque have restricted public data sharing. Relevant excerpts of the transcripts supporting each theme are included within the manuscript. Public sharing would not be consistent with the terms of participant consent. The full coding framework is available in S1 Table

## Acknowledgments

We thank the patients at Hospital Nacional de Coatepeque who generously shared their time and experiences during their hospitalization. We are grateful to the nursing staff of the orthopedic trauma wards for their assistance in identifying and screening eligible participants, and to hospital leadership for supporting this work.

## Supporting information

**S1 Text. Interview guide.** Semi-structured interview guide used for participant interviews.

**S1 Checklist. COREQ checklist.** Completed consolidated criteria for reporting qualitative research (COREQ) checklist.

**S1 Table. Codebook.** Codes generated during inductive coding of participant interviews and their mapping to preliminary thematic categories, which were subsequently consolidated into the five final themes.

